# KleTy: integrated typing scheme for core genome and plasmids reveals repeated emergence of multi-drug resistant epidemic lineages in Klebsiella worldwide

**DOI:** 10.1101/2024.04.16.24305880

**Authors:** Heng Li, Xiao Liu, Shengkai Li, Jie Rong, Shichang Xie, Yuan Gao, Ling Zhong, Quangui Jiang, Guilai Jiang, Yi Ren, Wanping Sun, Yuzhi Hong, Zhemin Zhou

**Author notes:** **Corresponding author:** Zhemin Zhou, Yuzhi Hong, Wanping Sun. Heng Li, Xiao Liu, Shengkai Li, and Jie Rong contributed equally to this work.

## Abstract

**Background:** Clinically important lineages in *Klebsiella*, especially those expressing multi-drug resistance (MDR), pose severe threats to public health worldwide. They arose from the co-evolution of the vertically inherited core genome and horizontal gene transfers by plasmids, which has not been systematically explored.

**Results:** We designed KleTy, which consists of dedicated typing schemes for both the core genome and plasmids in *Klebsiella*. We demonstrated the superior performance of KleTy over many state-of-the-art pipelines using both simulated and real data. We used both schemes to genotype 33,272 *Klebsiella* genomes, categorising them into 1,773 distinct populations and predicting the presence of 87,410 plasmids from 837 clusters (PCs). Notably, *Klebsiella* is the center of the plasmid-exchange network within Enterobacteriaceae. Our results associated the international emergence of prevalent *Klebsiella* populations with only four carbapenem-resistance (CR) PCs, two hypervirulent PCs, and two hvCR-PCs encoding both carbapenemase and hypervirulence. Furthermore, we observed the ongoing international emergence of *bla*_NDM_, accompanied by the replacement of the previously dominant population, *bla*_KPC_-encoding HC1360_8 (CC258), during 2003-2018, with the emerging *bla*_NDM_-encoding HC1360_3 (CC147) thereafter. Additionally, expansions of carbapenem-resistant *Klebsiella pneumoniae* (hvCRKP) were evidenced in both populations, driven by plasmids of MDR-hypervirulence convergences.

**Conclusions:** The study illuminates how the global genetic landscape of *Klebsiella* has been shaped by the co-evolution of both the core genome and the plasmids, underscoring the importance of surveillance and control of the dissemination of plasmids for curtailing the emergence of hvCRKPs.

## Background

Nosocomial infections by carbapenem-resistant *Klebsiella pneumoniae* (CRKP) that also express multidrug resistance (MDR), first reported in the 1990s, now have a worldwide distribution [1]. In particular, the emergence of hypervirulent CRKPs (hvCRKPs) in China, India, and many other countries has drawn special attention [2] due to their association with the increased prevalence of bloodstream infections [3]. To investigate infections and transmission of *K. pneumoniae*, a wide range of typing techniques have been utilized [4], including serotyping, pulsed-field gel electrophoresis (PFGE), and multi-locus sequence typing (MLST). Recently, cgMLST schemes for the *K. pneumoniae* Species Complex (KpSC) have been developed and hosted by Institut Pasteur (629 genes) [5], which also designed cgLINcodes to infer population structures. Clinical use of the cgMLST scheme, however, has been restricted by the requirement to upload nucleotide sequences into central databases, which can be difficult for clinicians and/or epidemiologists [6]. The process also risks data privacy. As a result, state-of-the-art pipelines, such as Kleborate [4], still rely on MLST and serotyping to characterize *Klebsiella* strains.

Mobile genetic elements harboring antimicrobial-resistant genes (ARGs) and virulence factors (VFs) have been regarded as one of the major driving factors behind the emergence of epidemic lineages in *K. pneumoniae* [7]. Notably, the recent emergence of ST11 hvCRKP strains in China has been associated with the acquisition of virulence factors (VFs) in the CRKPs [8], calling for systematic analyses of the complex interplay between the plasmids and the bacterial hosts. In addition, *K. pneumoniae* has long been regarded as the hub for inter-species horizontal genetic transfers (HGTs) of plasmids, especially among *Enterobacteriaceae* [9]. Investigation of the contextual genetic diversity of plasmids in *Klebsiella* will facilitate our understanding and control of the inter-species spreading of ARGs, which has been frequently associated with the plasmids [1]. Currently, low resolution limits state-of-the-art techniques, such as replicon typing and MOB typing [10], thereby highlighting the need for new algorithms that systematically predict and catalog plasmid content.

Here we describe an automatic pipeline, KleTy, that enables unified genotyping of both the core genome and the plasmids in all *Klebsiella* species. KleTy consists of three modules: 1) a population assignment module based on a novel dcgMLST+HierCC scheme, 2) a plasmid prediction module based on a novel plasmid clustering (PC) scheme, and 3) an ARG/VF prediction module. We showed that KleTy outperformed state-of-the-art pipelines in both population assignments and plasmid predictions based on the prediction of 1,773 natural populations and 87,410 plasmids in 33,272 *Klebsiella* genomes. This work expanded our understanding of the genetic diversity of plasmids in *Klebsiella* and the dynamics of predominant populations that are associated with the global elevation of hvCRKP over decades.

## Results

### A unified genotyping scheme for chromosome and plasmid in *Klebsiella*

Here we establish KleTy, a tool that rapidly genotypes core genomes and plasmids in *Klebsiella* with three modules (Fig. 1a). Based on KleTy, one can easily assign a given *Klebsiella* genome to a pre-defined population and identify clinically relevant genes and plasmids in minutes.

**Figure 1.**
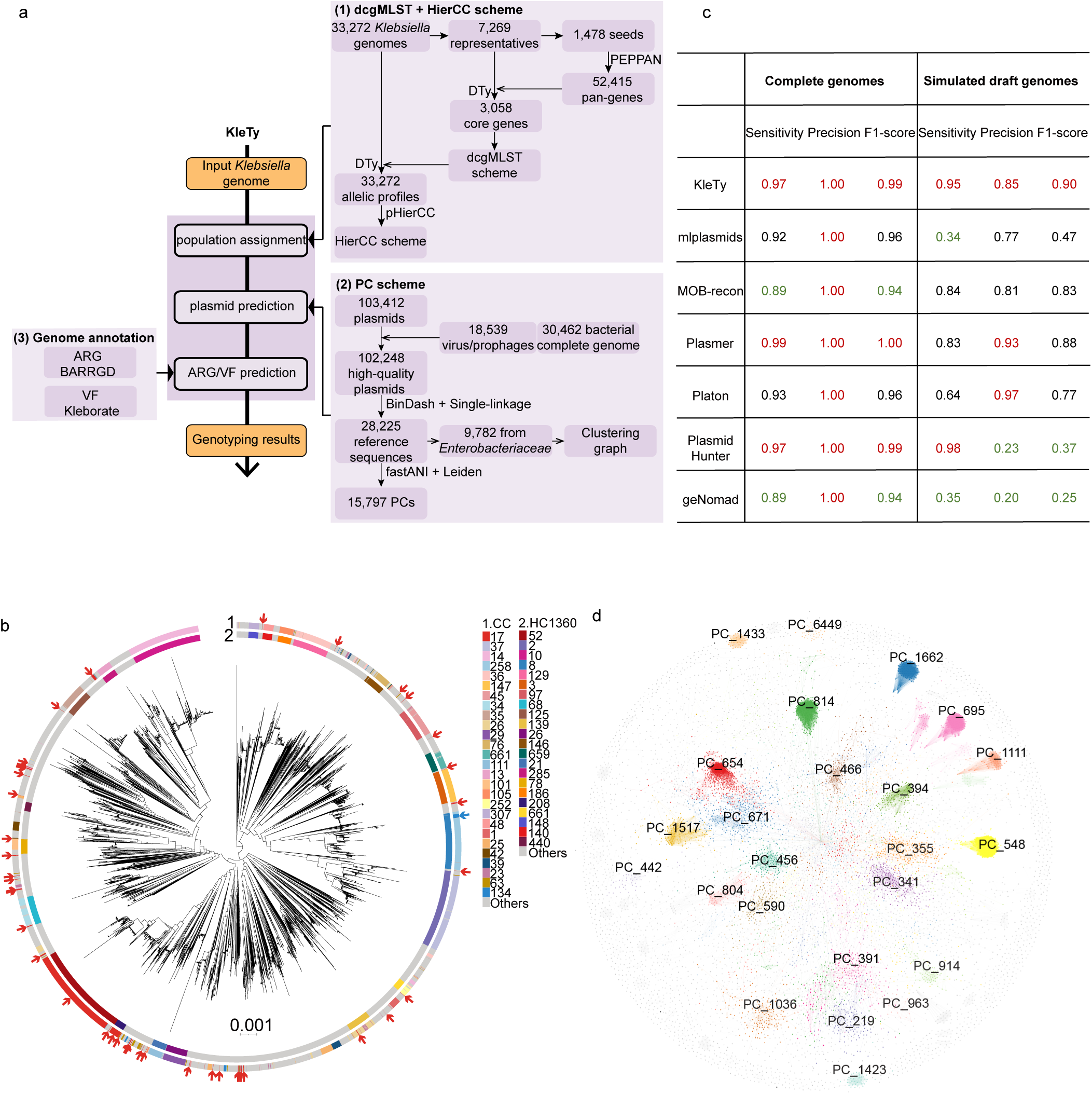
The workflow and evaluation results for the KleTy pipeline. (a) KleTy consists of three modules that (1) genotype each genome into a set of curated hierarchical clusters using the dcgMLST+HierCC scheme, (2) identify plasmids and assign each to one of the 15,826 clusters in the Plasmid Clustering (PC) scheme, and (3) predict ARGs and VFs based on curated gene sets. (b) Visualizing the HC1360 groups and the clonal complexes (CCs) in the species tree of 5109 *K. pneumoniae* genomes, which is a subtree of the genus-level supertree summarized from 3,058 gene trees (see Methods). The circular bars surrounding the tree showed (from outer to inner) the CC and the HC1360 assignments of each genome (inset Keys). Arrows show the genomes inaccurately assigned to CC17 (red) or CC134 (blue). (c) Comparisons of the plasmid predictions by KleTy, mlplasmids, MOB-recon, Plasmer, Platon, PlasmidHunter, and geNomad on benchmark datasets of 1,271 complete *Klebsiella* genomes (columns 1-3), and 100 simulated draft genomes (columns 4-6). The simulated draft genomes were assembled from simulated short reads by wgsim based on randomly selected complete genomes. (d) The similarity network of 9,782 reference plasmids from *Enterobacteriaceae*. Plasmids (nodes) from the top 30 most abundant PCs were color-coded.

### Population assignment module based on dcgMLST+HierCC scheme

A total of 3,058 core genes were selected to establish a distributed cgMLST (dcgMLST) scheme [6] from 33,272 *Klebsiella* genomes. Furthermore, we evaluated a series of single-linkage clustering results with varied allelic differences and identified an optimal level of HC1360 for *Klebsiella*, which subdivided all 33,272 genomes into 1,773 distinct clusters that broadly represented natural populations (Table S1). These HC1360 clusters were well matched to the clonal complexes (CCs) in the 7-gene MLST scheme with an adjusted Rand index of 0.967, leaving only a few discrepancies (Table S2).

To investigate discrepancies between HC1360s and CCs, a species tree of *Klebsiella* was constructed by combining 3,058 trees of the core genes using a divide-and-conquer strategy [11]. After mapping both HC1360s and CCs onto the species tree (Fig. 1b), we found that all HC1360s formed monophyletic clusters in the species tree, whereas 16.8% of CCs mistakenly generated paraphyletic groups. For example, CC17 was found in 42 monophyletic clades across the tree and CC134 emerged as a sub-clade within the CC258 clade, thereby underscoring the superior performance of HC1360s over CCs.

### Plasmid prediction module based on pre-curated plasmid clusters (PCs)

We retrieved all 103,412 complete plasmids hosted by >2,400 bacterial species in GenBank (as of March 2023) to capture all known genetic diversity of the plasmids. After removing potential mislabeling chromosomal or viral DNAs, the remaining high-quality plasmid genomes were grouped into 15,797 plasmid clusters (PCs) using the Leiden algorithm [12] with an average nucleotide identity of ≥90% and alignment coverage of ≥50% (Fig. 1d; see Methods). Furthermore, we constructed a plasmid-exchange network of *Enterobacteriaceae* by bridging each PC with its associated genera (Fig. 2a) and found that 68% of the *Klebsiella*-harbored PCs were also detected in other genera including *Escherichia*, *Enterobacter*, and *Salmonella*. This makes *Klebsiella*, which had the greatest eigenvector centralities of 1.0, the predominant center of the plasmid-exchange network (Fig. 2b, Table S3).

**Figure 2.**
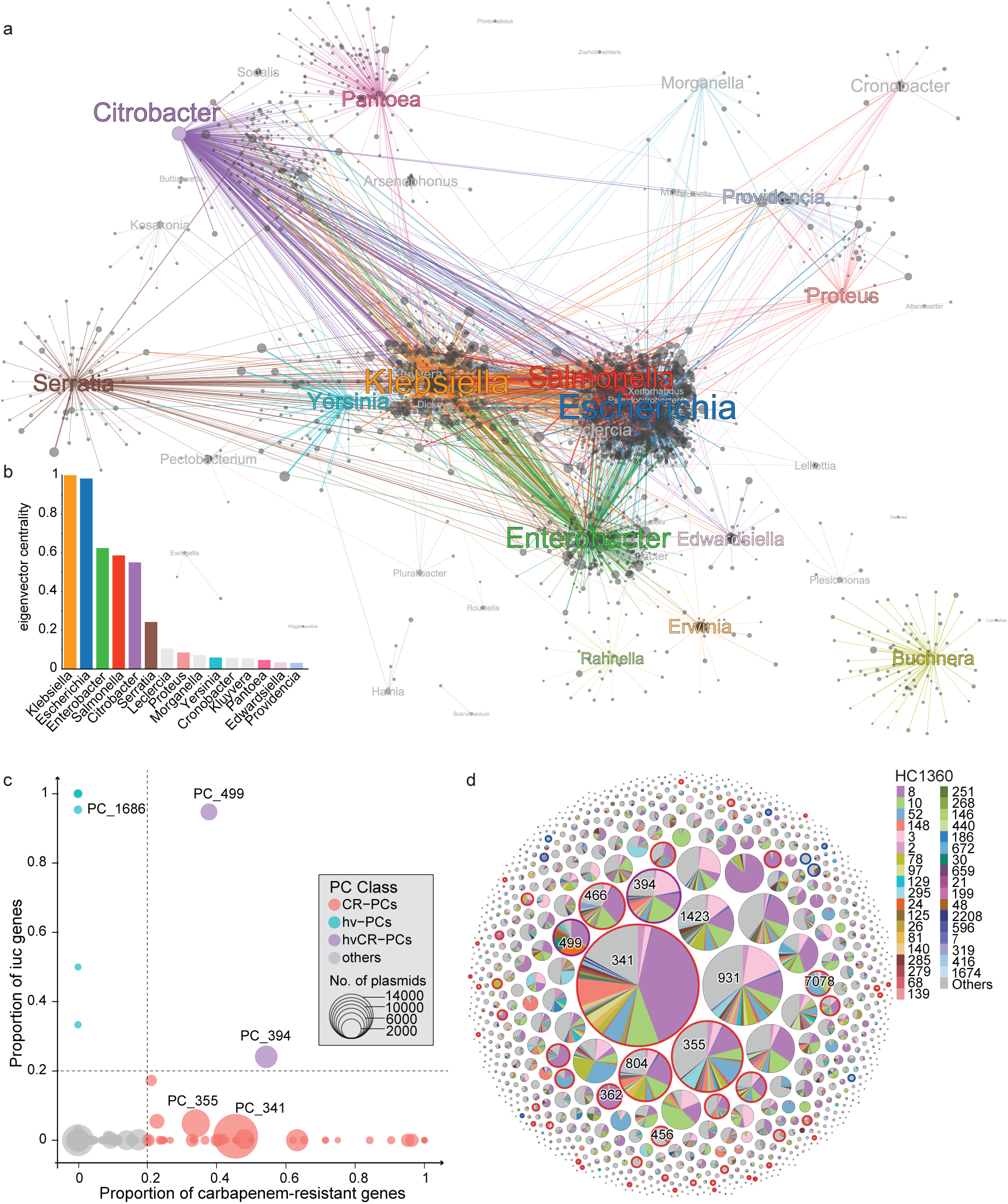
The host association and genetic characteristics of the PCs. (a) A graph of the plasmid-exchange network in *Enterobacteriaceae*. Nodes show the plasmids (grey) and genera of the hosts (colored) and the colored edges show the presence of the plasmids in the corresponding hosts. (b) Histogram of the eigenvector centralities of genera in the network in part a. Additional metrics used to quantify the importance of nodes in the plasmid-exchange network are shown in Table S3. (c) Scatter plot of the average carriage of carbapenemase genes (X-axis) and the *iuc* gene (Y-axis) in each PC. Each circle represents a PC and is sized proportional to the number of associated plasmids and color-coded as in the Key. The two dashed lines show the criteria for assigning PCs as CR-PCs (≥0.2 carbapenemases), hvPCs (≥0.2 *iuc*), or hvCR-PCs. (d) Bubble plot of the HC1360 distribution for each PC. Each bubble indicates a PC and is sized relative to the associated plasmids. The piechart in each bubble shows the proportional presence of the PC in different HC1360s. The halos surrounding some bubbles indicate that these PCs belong to one of the CR-PCs (red), hvPCs (blue), or hvCR-PCs (purple), as defined in part C.

We designed an algorithm as the plasmid module in KleTy for predicting PCs and compared its performance with six existing algorithms, including mlplasmids [13], MOB-recon [10], Plasmer [14], Platon [15], PlasmidHunter [16], and recently published geNomad [17] based on two datasets (Table S4) of (a) 1,271 complete *Klebsiella* genomes containing 4,796 plasmids, and (b) 100 simulated draft assemblies containing 6,597 plasmid-originated contigs (see Methods). KleTy’s results had high accuracies with F1 scores of 0.99, and 0.90 in each of the two datasets, respectively. In contrast, while the other six tools performed decently with complete genomes (F1 scores of 0.94-1.00), their performances dropped substantially with the draft genomes, with F1 scores of only 0.47, 0.83, 0.88, 0.77, 0.37, and 0.25, for results from mlplasmids, MOB-recon, Plasmer, Platon, PlasmidHunter, and geNomad, respectively (Fig. 1c).

### ARG/VF prediction Module

To evaluate the clinical importance of predicted populations and plasmids, we employed the third module that predicts ARGs and VFs based on genomic data by enrolling reference genes in the “Bacterial Antimicrobial Resistance Reference Gene Database” (https://www.ncbi.nlm.nih.gov/bioproject/PRJNA313047) and “Kleborate” (https://github.com/klebgenomics/Kleborate/tree/main/kleborate/data). KleTy also uses the metadata in the ResFams database (https://www.dantaslab.org/resfams) for the recognition of ESBL and inhibitor-resistant variants of β-lactamases. Additionally, it ignores mutations in the *pmrB*, *oqxABR*, *ramR*, *uhpT*, and *fosA* genes due to the lack of evidence linking them to resistance in *Klebsiella* [18].

### Genetic landscape of populations and plasmids in *Klebsiella*

We identified 1,773 HC1360 populations in *Klebsiella* based on all 33,272 publicly available genomes (Fig. 3a), making it slightly more divergent than *Escherichia/Shigella*, which has 1,379 HC1100 populations [11]. Over 70% of the HC1360s fell within only three species of *K. pneumon*iae (43%, 757), *Klebsiella variicola subsp. variicola* (17%, 299), and *Klebsiella quasipneumoniae subsp. similipneumoniae* (11%, 193) (Table S5). Notably, nearly half of the *Klebsiella* genomes were from one of the five predominant populations of HC1360_8 (CC258; 8587 strains), HC1360_10 (CC14; 2403), HC1360_52 (CC17; 1714), HC1360_148 (CC307; 1645), and HC1360_3 (CC147; 1493) (Fig. 3d). HC1360_8 (CC258) is the primary source of *Klebsiella* in China, Israel, the US, and countries in South America and Europe; HC1360_10 (CC14) predominates Saudi Arabia and many countries in South/Southeast Asia (Fig. 3c). HC1360_148 (CC307) and HC1360_52 (CC17) were prevalent in France and Thailand, respectively. None of the predominant HC1360s has been frequently found in North Europe or Africa, indicating their distinct epidemiological patterns.

**Figure 3.**
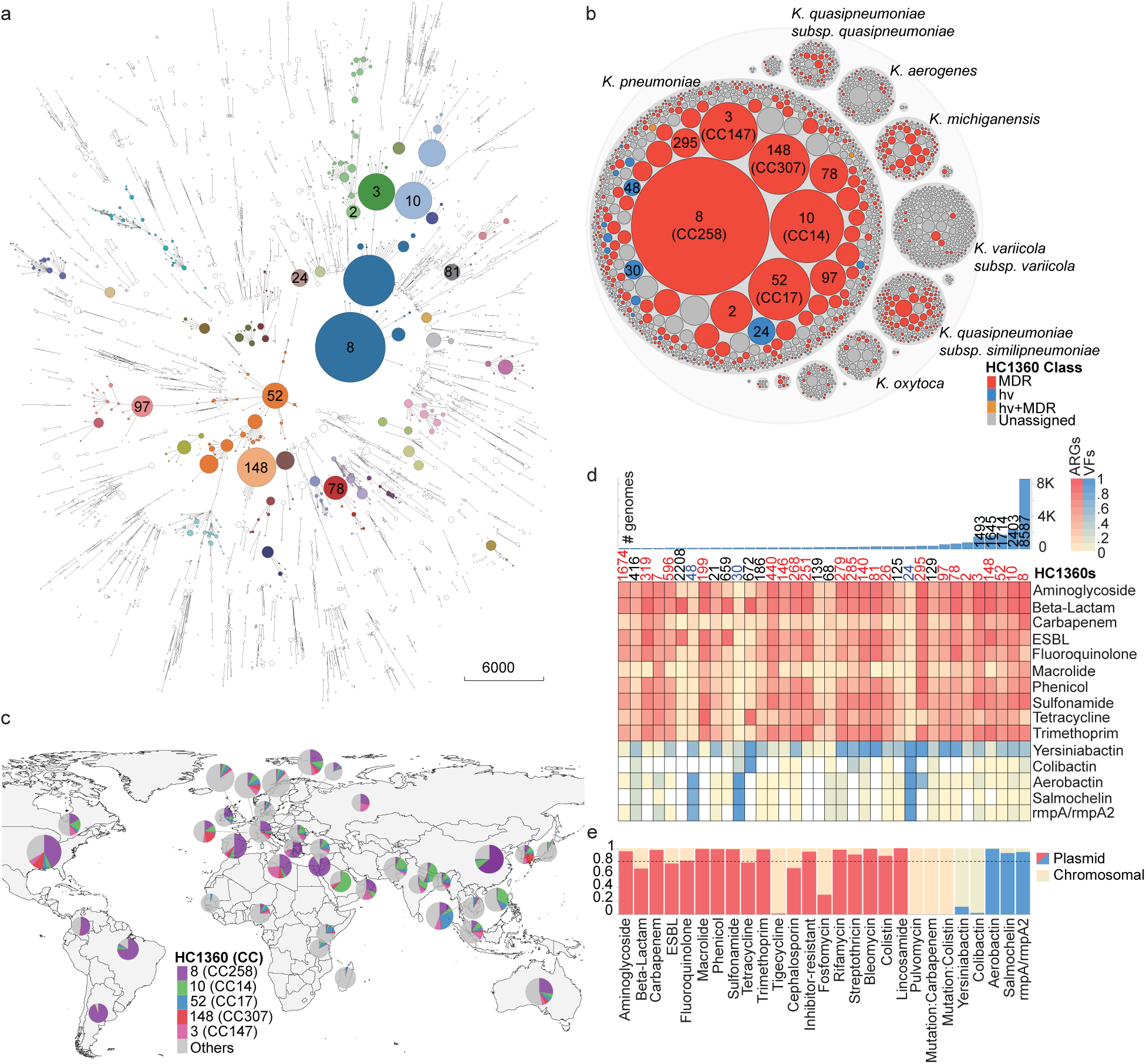
The population structure and geographical distribution of the *Klebsiella* genomes. (a) A minimum spanning tree of 33,272 *Klebsiella* genomes based on their dcgMLST profiles. Branches with <400 allelic distances are collapsed. The HC1360 populations with ≥100 genomes are color-coded and some are also labeled. Only branches with <3035 allelic distances are shown by clarity. (b) Hierarchical bubble plot for the population structure of the *Klebsiella* genus. The bubbles indicate the genus, species, and HC1360 populations in *Klebsiella*. Some HC1360s are color-coded because the majority of the genomes in them are MDR (red), hypervirulence (blue), or both (orange). (c) Global distribution of the five major HC1360 populations in countries with ≥100 genomes visualized using D3.js. (d) The heat plot of the prevalence of antimicrobial-resistant genes (ARGs; red) and hypervirulence factors (VFs; blue) in the top 36 HC1360 populations. The histogram on the top shows the numbers of genomes in the HC1360 populations. (e) The predicted percentages of each of the ARGs and VFs from the plasmids. The dotted line indicates 80% of plasmid origins.

KleTy also predicted the presence of 87,410 plasmids from 837 PCs in *Klebsiella* (Table S6). Of the identified PCs, 38% (314/837) were novel for *Klebsiella*, having been previously found only in other genera, such as *Enterococcus*, *Escherichia*, *Staphylococcus*, *Acinetobacter*, *Bacillus*, *Neisseria*, *Enterobacter*, and *Salmonella* (Table S7). KleTy assigned Incompatibility (Inc) types to ∼78% (68,103/87,410) of the plasmids, with ColRNAI, IncFIB, and IncFII being the most frequent (Fig. S1a). It also assigned 46,715 (53%) plasmids to MOB types, with MOB_F_, MOB_P_, and MOB_H_ being the most frequent (Fig. S1b). There was minimal association between typing schemes, with most PCs linking to multiple Inc and MOB types. In summary, our findings substantially expand our understanding of the genetic diversity in *Klebsiella*.

### Emergence of MDR HC1360_8 (CC258) population with *bla*_KPC_-carrying plasmids

Approximately 29% (513/1,773) of the HC1360s from 13 *Klebsiella* species are MDR, with ≥3 ARGs on top of the intrinsic genes of *bla*_SHV_, *bla*_LEN_, *bla*_OKP_, and *oqx*. Half (265/513) of these MDR HC1360s fell among *K. pneumoniae*, followed by *K. quasipneumoniae subsp. similipneumoniae* (82) and *K. michiganensis* (58) (Fig. 3b).

Nearly half (47%; 15,509/33,272) of the *Klebsiella* genomes exhibited carbapenem resistance (CRKP) due to the acquisition of carbapenemases (Fig. 4a). We found 50 carbapenem-resistant PCs (CR-PCs) that each has a ≥20% carbapenemase carriage rate (Fig. 2c). CR-PCs accounted for four of the six most abundant plasmids in *Klebsiella* (Fig. S2), including PC_341 (37% *bla*_KPC_, 88% IncFIB/IncFII), PC_355 (17% *bla*_KPC_, 60% IncFIB/IncFII), PC_394 (34% *bla*_NDM_, 80% IncFIB/IncHI1B), and PC_804 (51% *bla*_OXA_, 99% IncL/M). Most of these CR-PCs were each associated with three or more categories of carbapenemases (Fig. S2), as previously reported [19]. Meanwhile, geographically restricted CR-PCs also exhibited almost exclusive association with only one category of carbapenemase (Fig. 4b), such as the *bla*_KPC-2_-encoding PC_362 and PC_499 in China and *bla*_KPC-3_-encoding PC_396 and PC_1671 in the US.

**Figure 4.**
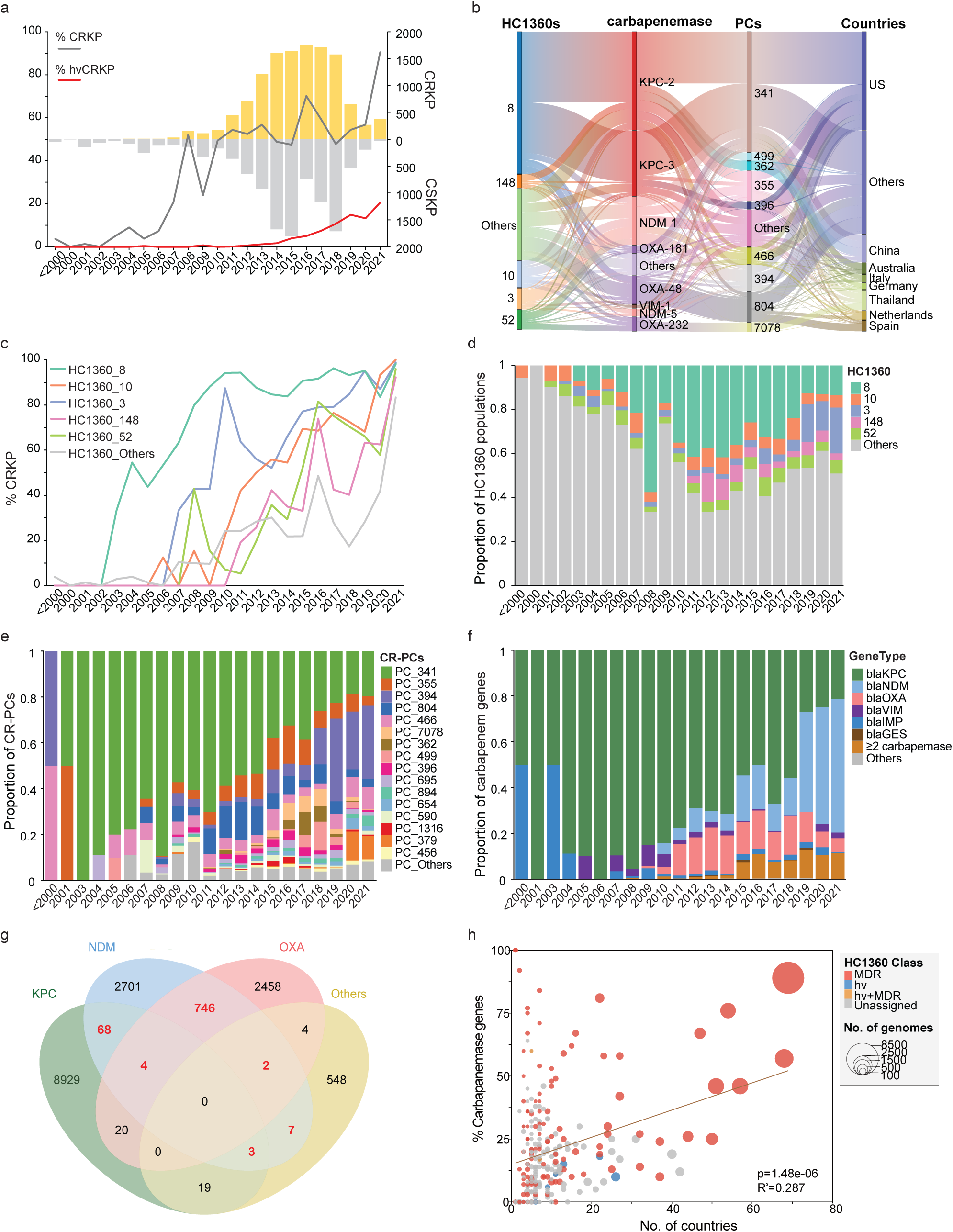
The prevalences and genetic characteristics of carbapenemase-carrying *Klebsiella* strains. (a) The annual frequencies of CRKP (yellow) and carbapenam-sensitive *Klebsiella* (CSKP; grey) in the 23,868 genomes with known isolation years. The relative percentages of the CRKP (dark grey) and hvCRKP (red) in each year are also shown as curves. All strains isolated before 2000 are assigned to one bin for clarity. (b) The Sankey diagram shows the relationships between the HC1360 populations, carbapenemases, PCs, and the countries of the CRKPs. (c) The percentages of CRKPs per year for each of the five predominant HC1360 populations and the remaining. (d-f) The relative proportions of the predominant HC1360 populations (d), CR-PCs (e), and carbapenamases (f) per year. (g) Venn diagram of the carbapenemase profiles of the CRKPs. Each oval shows the carriages of one of the *bla*_KPC_, *bla*_NDM_, *bla*_OXA_, and other carbapenamases. The overlapping regions show genomes that each encode two or three carbapenemase categories simultaneously. The red numbers show the multi-carriage of both *bla*_NDM_ and other carbapenamases. (h) The bubble plot of the country distributions (X-axis) and percentage of CRKPs (Y-axis) for each HC1360 population. The circles were sized relative to the number of genomes and color-coded according to the Key. The black line shows a positive correlation between the CRKP percentages and the country distributions of HC1360s by the linear regression (Pearson’s R^2^ = 0.287, p=1.48e-6). We also spotted a similar positive correlation between the CRKP percentages and the numbers of genomes of the HC1360s (R^2^ = 0.294, p= 7.55e-07).

Very few of the strains collected before 2003 are CRKPs (1.4%, 4/290). As of 2003, one HC1360_8 strain acquired a *bla*_KPC-3_-encoding PC_341 plasmid, which gradually increased its frequency in the HC1360_8 (CC258) population over the next five years, reaching >80% carriage in 2008. Meanwhile, the isolation frequencies of HC1360_8 (CC258) strains also increased rapidly, accounting for 58% of *Klebsiella* and 89% of CRKPs in 2008 (Fig. 4c, d). HC1360_8 strains isolated early were mostly ST258 from the US, but the Chinese *bla*_KPC-2_-producing ST11 strains, first reported in 2007 [20], later also became one of the primary sources [8]. Other *Klebsiella* populations also accumulated carbapenemases during 2006-2011. For example, the *bla*_OXA-48_ carried by PC_804 accounted for ∼12% of the isolates between 2011 and 2014 (Fig. 4b, e). However, the *bla*_KPC_-encoding HC1360_8 kept being the most prevalent CRKP population, accounting for ∼30% of annual *Klebsiella* isolates until 2019 (Fig. 4d), when it was surpassed by the *bla*_NDM_-encoding HC1360_3 (CC147).

### Emergence of HC1360_3 (CC147) with plasmid-driven *bla*_NDM_-hypervirulence convergence

HC1360_3 (CC147) was the most abundant population after 2019, accounting for 13-21% of the *Klebsiella* strains (Fig. 4d). It exhibited a diverse carbapenemase profile of *bla*_NDM-1_ (53%, 606/1138), *bla*_NDM-5_ (7%, 77), *bla*_OXA-48_ (10%, 111), and *bla*_KPC-2_ (6%, 68). We found an increase of *bla*_NDM_ in HC1360_3 up to 89% (317/356) during its upsurge since 2019, indicating a possible association between *bla*_NDM_ and the population expansion (Fig. 4f, Fig. S3). In addition, the recent growth of *bla*_NDM_ carriage rates in less common populations was also observed. Over 70% of strains in HC1360s with <1000 strains carry *bla*_NDM_ after 2020, resulting in the general elevation of carbapenem resistance in *Klebsiella* to 90% by 2021. Moreover, *bla*_NDM_ contributed to 94% of the 883 multi-carbapenemase strains, which each carried two or more carbapenemases, further underscoring its complicated genetics (Fig. 4g).

Virulence also contributes to the epidemiology of *Klebsiella*. Hypervirulence in *Klebsiella* has been associated with the presence of five virulence loci, especially *iuc*, which encodes the siderophores aerobactin. The hvKPs and the CRKPs normally fall into different HC1360s (Fig. 3d) and are associated with the acquisition of different plasmids (Fig. 2d). However, some HC1360s have been reported to experience AMR-virulence convergence, resulting in severe disease outbreaks [8,21]. Using both *iuc* and carbapenemase genes as markers, we observed a steady increase of hvCRKP frequencies over time, from 4.5% before 2019 to 15.8% currently (Fig. 4a). This increase has been associated with convergence or conjugation of hv-and ARG-carrying plasmids [22].

Two hvCR PCs had high levels of both *iuc* and carbapenemase genes: PC_499 (94.7% *iuc*, 37.8% carbapenemase, 89% IncFIB/IncHI1B) and PC_394 (24.1% *iuc*, 54.3% carbapenemase, 80% IncFIB/IncHI1B) (Fig. 2c). PC_499 resulted from the conjugation of pLVPK (also in PC_499), the most abundant hv-PC in *Klebsiella*, and the *bla*_KPC-2_-carrying plasmids in China (Fig. 5a). It has been exclusively associated with the recent emergence of the ST11-K64 hvCRKP clone in China and rarely found elsewhere. Through a phylogenetic analysis of 2,530 ST11 genomes from China, we observed that >90% of the ST11-K64 hvCRKPs fell into one monophyletic cluster that was associated with a cluster of PC_499 hvCR plasmids (Fig. 5b). The hvCRKPs were much fewer outside this cluster and were associated with acquisitions of multiple plasmids.

**Figure 5.**
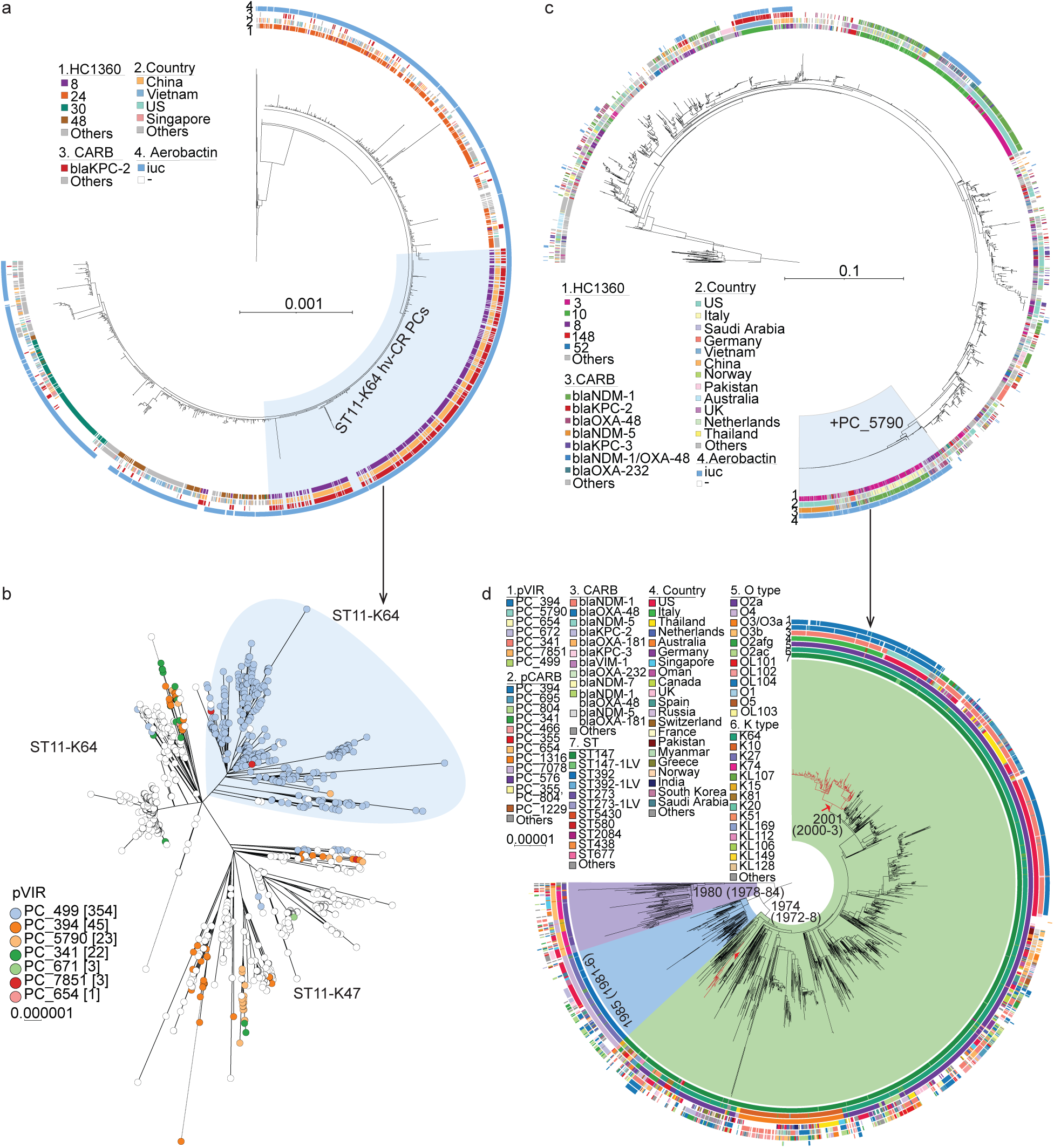
The maximum-likelihood phylogeny of the major hvCRKP HC1360 populations and their associated hvCR-PCs. (a) The phylogeny of 1,098 PC_499 plasmids in this study and 344 public PC_499 plasmids. The hvCR-PCs responsible for the majority of the hvCRKP in ST11-K64 in part B are highlighted in light blue. (b) The phylogeny of 1,114 ST11 genomes that are from China or fall within the same clade of the Chinese strains. The shape in light blue highlights a cluster of ST11-K64 strains that are mostly hvCRKPs due to the acquisition of the hvCR PC_499 plasmids in part A. (c) The phylogeny of 2,608 PC_394 plasmids in this study and 378 public PC_394 plasmids. The hvCR-PCs responsible for the majority of the hvCRKPs in ST147 in part d resulted from a conjugation of the plasmids in PC_5790 and are highlighted in light blue. (d) The phylogeny of 1,493 global HC1360_3 (CC147) genomes. The three MLST STs associated with HC1360_3 (CC147) are shown in colored arcs and the hvCRKP clades are highlighted in red, carrying the hvCR PC_394 plasmids in part c. The circular bars in parts a, c, and d show metadata associated with the plasmids or genomes, as in the Keys.

PC_394 has been associated with the currently emerging HC1360_3 (CC147) [23] and causing disease outbreaks internationally [24,25] (Fig. 5c). It was formed by conjugation of the carbapenemase-carrying processor in PC_394 and the hypervirulence PC_5790. To investigate the population dynamics of HC1360_3 (CC147) and PC_394, we reconstructed a maximum-likelihood phylogeny based on 25,144 SNPs in the non-repetitive, non-recombinant core genome of 1,493 international HC1360_3 strains (Fig. 5d) and estimated its date of origin and geographic transmission (Fig. S4). The most recent common ancestor (MRCA) of HC1360_3 was estimated to be present before 1947 (CI95%: 1945-1954) in the US, and it later diverged there into three lineages that broadly consisted of strains from ST147 (1,244 strains), ST392 (139), and ST273 (90). The ST147 lineage likely emerged before 1974 (CI95%: 1972-1978) and was gradually transmitted into ≥48 countries. A transition of primary carbapenemase from *bla*_OXA_ or *bla*_KPC_ in early strains to *bla*_NDM_ encoded by PC_394 or PC_695 after 2017 was observed (Fig. 5d). Furthermore, we predicted many AMR-virulence convergences along the phylogeny of PC_394, including one large cluster resulting from conjugation with PC_5790 plasmids (Fig. 5c). The resulting hvCR plasmids were independently acquired by HC1360_3 (CC147) to form hvCRKPs ≥9 times. Two of the resulting hvCRKP clusters have been circulating for ≥10 years, one responsible for repetitive infections in Russia [26], and the other for outbreaks in both Italy[24] and the US[25].

### Colistin resistance plasmids promote specific gene transmission across regions and hosts

Mobilized colistin resistance gene (*mcr*) resulted in reduced susceptibility of colistin, further limiting the treatment options [27]. We identified a total of 23 *mcr*-carrying PCs in *Klebsiella*. Most of these PCs were low in amount or had low *mcr* carriages, except for two, PC_456 and PC_1293, which accounted for 70% (205/292) of *mcr* in *Klebsiella*.

The IncHI2A PC_456 carries an average of eight ARGs per plasmid and was associated with two *mcr* variants, *mcr*-1 (6%) and *mcr*-9 (51%), each by plasmids from a different phylogenetic clade (Fig. 6a). The *mcr*-1-carrying clade consists of plasmids from Asian *Klebsiella* strains, as well as many from *Salmonella* and *Escherichia*. Meanwhile, the *mcr*-9-carrying clade consists of primarily Euroamerican strains, plus plasmids from *Enterobacter*, *Citrobacter*, and *Escherichia*. These findings reflected the presence of two dynamic plasmid pools in PC_456 each associated with different *Enterobacteriaceae* spp. In contrast, the IncX4 PC_1293 is rarely associated with ARGs other than *mcr*. It has been primarily found in *Klebsiella*, *Escherichia*, and *Salmonella*. All *mcr*-1-carrying plasmids fell into a genetically closely related cluster in the phylogeny, which accounts for 50% of the PC_1293 plasmids (Fig. 6b).

**Figure 6.**
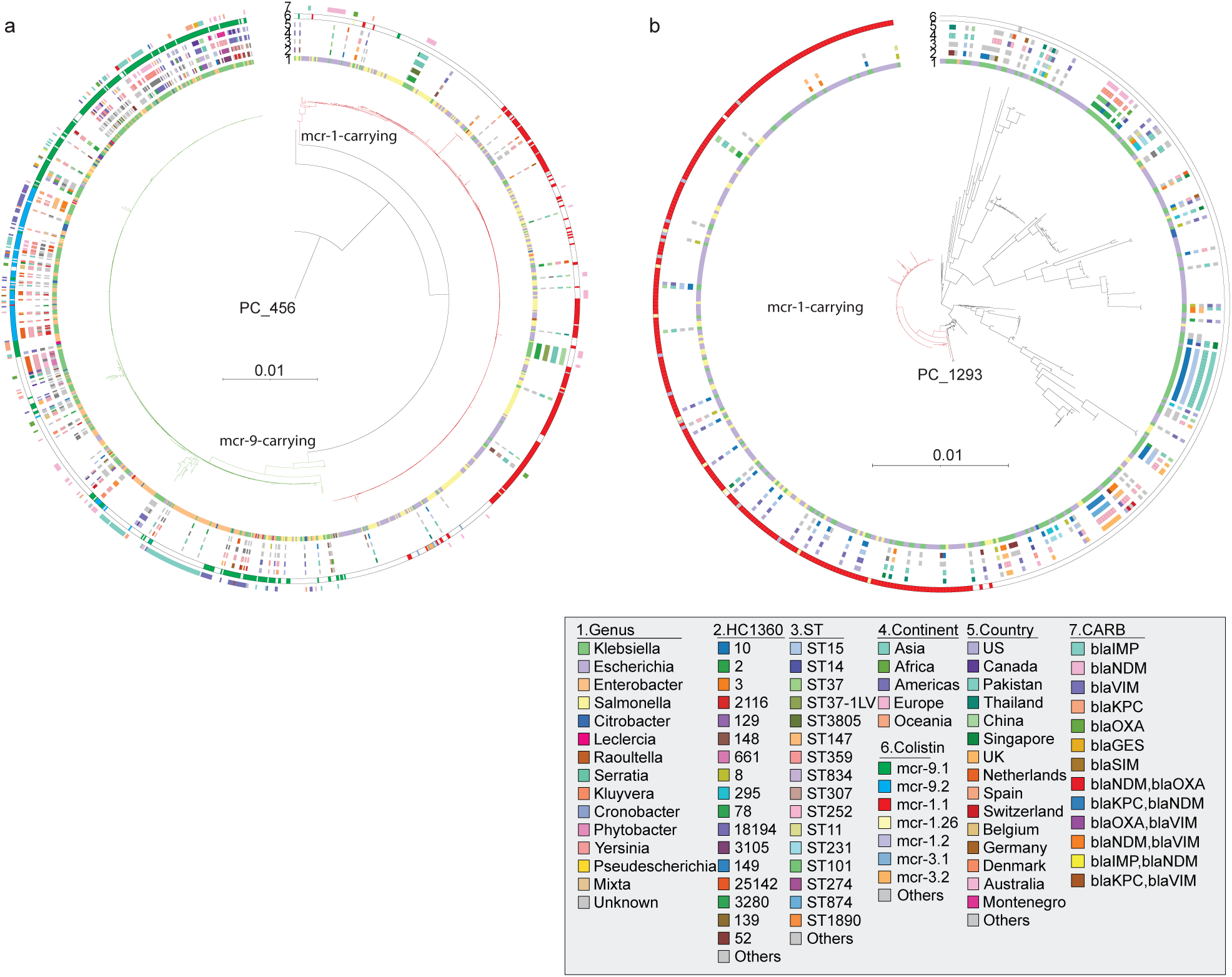
The maximum-likelihood phylogenies of *mcr*-carrying plasmids. (a) The phylogeny of 304 PC_456 plasmids in this study and 971 public PC_456 plasmids. Red and green branches are each associated with *mcr*-1 or *mcr*-9 genes, respectively. (b) The phylogeny of 149 PC_1293 plasmids in this study and 379 public PC_1293 plasmids. Red branches indicate plasmids carrying the mcr-1 gene. CARB: carbapenemase, ST: sequence type.

## Discussion

*Klebsiella* lineages that are clinically significant, particularly those that exhibit multi-drug resistance, arise from the co-evolution of the genetically conserved core genome and the highly variable, accessory genes that have been horizontally transmitted. Here we describe KleTy, an integrated tool that offers high-resolution genotyping solutions for both the core genome and plasmids based on the dcgMLST+HierCC and PC schemes, respectively.

CgMLST schemes have been widely applied to hundreds of bacterial pathogens, including *K. pneumoniae*, for tracking disease outbreaks and transmission chains [5]. However, its reliance on central databases has become a point of failure of the system and has also raised concerns regarding data privacy [6]. Here we adopted a dcgMLST scheme for *Klebsiella*, which was proposed recently [6] to overcome the above problems by implementing MD5 hash for allele designations without a central database. Furthermore, based on the dcgMLST types, we assigned each *Klebsiella* genome to one of the HC1360 clusters that represent genetically separate populations. These populations previously have also been approximated as clonal complexes (CCs) using the legacy MLST scheme, but we showed that the HC1360 clusters are more compatible with the *Klebsiella* phylogeny than the CCs (Fig. 1b). Additionally, the CC designations are unstable and may merge when adding in new strains [28]. The HierCC scheme overcomes the problem by implementing an algorithm that ensures static clustering assignments [29].

Plasmids have long been regarded as a “black hole” in phylogenetic research due to their highly variable gene contents and extensive HGTs across bacterial hosts [19]. KleTy managed to accurately predict plasmids and PCs based on a comprehensive reference dataset compiled from >100K existing plasmids. We demonstrated the superior performance of KleTy over six state-of-the-art pipelines of mlplasmids, MOB-recon, Plasmer, Platon, PlasmidHunter, and geNomad, in both complete and draft assemblies (Fig. 1c). Notably, all seven pipelines could be broadly separated by their algorithms into three categories: (1) similarity-based, which includes KleTy, MOB-recon, and Platon; (2) Kmer-based, which includes mlplasmids and Plasmer; (3) machine-learning based, which includes PlasmidHunter and geNomad. While the greatest sensitivity was found for results from PlasmidHunter, both machine-learning-based algorithms suffered from low precision of only 0.2-0.23 for the draft assemblies. In contrast, reduced sensitivity was found on the draft assemblies for the Kmer-based algorithms, especially mlplasmids which had a sensitivity of 0.34. The similarity-based algorithms, in general, had balanced sensitivities and precisions, and their performance was largely influenced by the quality of the associated databases. This indicates that the applications of machine-learning algorithms in detecting mobile genetic elements, such as plasmids, are still in need of further development.

We also used this massive PC dataset to investigate the dynamics of inter-species plasmid transfer among *Enterobacteriaceae*, which has been associated with the spread of ARGs [19] including carbapenemase and *mcr* genes (Fig. 2a). We demonstrated the pivotal role of *Klebsiella* and *Escherichia* in the plasmid-exchange network. These two species have been frequently associated with the worldwide dissemination of ARG-carrying plasmids, facilitating their transmission among other more virulent pathogens, such as *Salmonella enterica* and *Vibrio cholerae* [30,31]. Moreover, we showed that the plasmid-exchange rate has been drastically underestimated due to a lack of understanding of the genetic landscape of plasmids. KleTy identified 314 new PCs that have not been previously reported in *Klebsiella*, reflecting HGTs from/to other species in *Enterobacteriaceae* or even other families. Applying the PC module to other bacterial species in the future would allow a more systematic evaluation of the role of plasmid exchange for the cross-species spread of ARGs and other genes.

Notably, we found a lack of association between both Inc and MOB types and the PCs, consistent with previous reports [32]. Genes responsible for these traditional typings might have been horizontally transferred between different PCs, reflecting a new level of complexity. Intriguingly, among the 453 PCs that have been found between different species, 68% (308) have not been associated with MOB genes, which may have reflected a loss of MOB genes after the HGT. Alternatively, these plasmids might be hitchhikers that transferred together with the MOB-encoding plasmids in the same bacterial hosts [33].

Our analysis demonstrated the importance of chromosome-plasmid co-evolution in the formation of MDR epidemic lineages. Over 1/3 of the HC1360 populations in *Klebsiella* are MDR, mostly driven by plasmid-oriented ARGs (Fig. 3b, e). Notably, we identified 119 CRKP HC1360s that exhibited high levels (>80%) of carbapenem resistance and demonstrated positive correlations (Fig. 4h) between the carbapenem resistance of the HC1360s and the isolation frequencies (*P*<0.001) and geographic distributions (*P*<0.001).

It has also been clear that none of the chromosome, plasmid, or carbapenemase genes guarantees the success of a population. We found only weak associations between *Klebsiella* population and carbapenemase (Cramer’s V (CV): 0.37), carbapenemase and CR-PCs (CV: 0.29), and population and CR-PCs (CV: 0.45), indicating highly dynamic HGTs at all three levels. Even the five predominant populations are each associated with multiple carbapenemase genes and CR-PCs (Fig. 4b), and so are the five predominant PCs. Furthermore, while HC1360_8 (CC258) emerged after its acquisition of *bla*_KPC_-carrying PC_341, the same plasmid has also been found in 155 other HC1360s, most of which had very few clinical isolates. Similarly, *bla*_NDM_-carrying PC_394 was acquired by 99 HC1360s and resulted in the rise of only HC1360_3 (CC147) (Fig. 4). Moreover, *bla*_NDM_ genes spread along in the “minority” HC1360s independent of the selective advantages in the hosts. All these findings demonstrate the influence of co-evolution in the emergence of *Klebsiella* populations: only those that had selective advantages in both chromosome and plasmids demonstrate prevalence.

The plasmid-mediated colistin resistance genes were first discovered in 2015 [27], posing a significant threat due to their potential for rapid spread of colistin resistance. We associated 70% of the mcr genes in *Klebsiella* to only two PCs of PC_456 (IncHI2A) and PC_1293 (IncX4) (Fig. 6). PC_456 exhibited a broad host range and carried both *mcr*-1 and *mcr*-9. In contrast, PC_1293 exclusively carried the *mcr*-1 genes and was found in a narrower range of hosts. These findings highlighted the fact that the characteristics of the plasmids pose a strong influence on the spreading potential and destiny of its associated ARGs.

We revealed the emerging and global predominance of the HC1360_8 (CC258) lineage during 2003-2018 (Fig. 4). Carbapenem resistance in HC1360_8 was primarily attributed to *bla*_KPC_ genes (87%), *bla*_OXA_ (8%), and *bla*_NDM_ (6%), each associated with a different panel of plasmids. Over 52% of *bla*_OXA_ in HC1360_8 was associated with PC_804, which was also the primary source of *bla*_OXA_ in other populations. Geographic specificity was found in HC1360_8 for *bla*_KPC_-carrying plasmids, which were PC_341 in Europe and the US and PC_499 and PC_362 in China. Finally, similar to other populations, *bla*_NDM_ in HC1360_8 was associated with >20 PCs with no clear geographical preference. These findings confirmed previous reports [34] and further revealed that HC1360_8 has not carried a different carbapenemase than other, less prevalent populations. Most of its success is likely due to other factors, including core gene variations that have not yet been systematically investigated.

The recent emergence of hvCRKP due to AMR-virulence convergence raises major concerns due to the high bloodstream infection rates and limited antimicrobial treatment options[3]. While the overall frequencies of AMR-virulence convergences remained low (4.5%), we spotted a rapid increase of hvCRKP strains in the past five years, up to ∼20% (Fig. 4a). This could be partially attributed to sampling bias, while there was a general trend of two emerging populations: (1) *bla*_KPC-2_-carrying ST11-K64 hvCRKPs in HC1360_8 and (2) the *bla*_NDM_-carrying hvCRKPs in HC1360_3 (CC147). ST11-K64 hvCRKP strains have resulted from a convergence of classical virulence plasmid, pLVPK (PC_499) with CR-PCs[35]. It was first reported in 2017 in Zhejiang[8] and later also found in almost all provinces in China as part of an ongoing clonal replacement[36]. Our phylogenetic reconstruction of PC_499 (Fig. 5a) revealed that most (>90%) of the *bla*_KPC_-virulence convergence in ST11-K64 hvCRKPs are associated with a narrow spectrum of PC_499 plasmids, indicating high genetic stability of the convergence. Notably, these hvCRKPs have been rarely found in other countries, indicating the presence of other, unknown factors that limit the spread of hvCRKP. High associations have been recognised between hvCRKPs and the virulence plasmid markers (*iuc*, *iro*, *rmpA*, *rmpA2*, *peg-344*) [37]. Here we selected *iuc* as the marker for hv-PCs because it has been well investigated and exhibited a direct association with sepsis, promoting the blood growth of bacteria by acquiring iron from transferrin [4].

We also uncovered the recent emergence of hvCRKPs in HC1360_3 (CC147). This new hvCRKP group is of particular concern because both the bacterial hosts and the associated plasmids have been widely reported in Asia, Europe, and the Americas for decades before the AMR-virulence convergences. Furthermore, the HC1360_3 hvCRKPs have been causing disease outbreaks in Russia, Italy, and the US [24–26]. Our results showed that all three outbreaks were associated with one genetically stable hvCR cluster in PC_369, and the resulting hvCRKPs likely have been endemic in each region for decades. These hvCRKPs likely have contributed to the recent emergence of HC1360_3, which demonstrated global prevalence in the past three years. Effective research and controls are urgently needed for this previously underestimated population.

We acknowledge the limitations of our study. The potential sampling bias and incompleteness of the metadata may have limited the ability to accurately determine the prevalence of HC1360s and plasmids in specific regions. Additionally, the reference plasmid database may introduce bias, particularly for plasmids of highly variable or unknown sequences.

## Conclusions

In summary, we investigated the genetic landscape of *Klebsiella*, demonstrating the role of chromosome-plasmid interactions in facilitating the dissemination of antimicrobial resistance and virulence genes. We revealed two sequential global pandemic populations, HC1360_8 (CC258) which was primarily associated with *bla*_KPC_-carrying PC_341, and HC1360_3 (CC147) which has *bla*_NDM_-carrying PC_394. An ongoing expansion of carbapenemase-hypervirulence convergences was reported in both populations, underscoring the importance of understanding the association between plasmids and specific populations and genes, prompting monitoring of plasmids for effective prevention and control of serious infections caused by *K. pneumoniae*.

## Methods

### Genome sequence collection

To capture the broadest possible diversity of *Klebsiella* populations, we established a comprehensive genomic dataset of *Klebsiella* that consists of a total of 33,272 assemblies and short reads retrieved from GenBank (as of Aug. 2022, Table S8). All short reads were assembled into draft genomes using EToKi [38]. These genomes represent isolates collected between 1886 and 2022 from 94 different countries, with the majority from the Americas (27.8%, n=9,243), Europe (25.2%, n=8383), and Asia (20.5%, n=6815). The STs, ARG/VF profiles, and capsular types of each genome were predicted using Kleborate v2.3.2[4], and the genes were predicted and annotated using Prokka[39].

### Establishments of the dcgMLST+ HierCC schemes

Construction of the dcgMLST scheme consisted of three stages (Fig. 1a). First, to reduce genetic redundancy due to the overrepresentation of genetically nearly identical strains in *Klebsiella*, we employed kssd [40] to estimate the pairwise genetic distances of all 33,272 genomes, and separated them into single-linkage clusters of ≥99.8% identities. One genome with the greatest N50 value was chosen for each cluster and subjected to quality checking using FetchMG [41]. A total of 7,269 genomes were kept because they each carried ≥37/40 single-copy core genes (SCGs), forming the representative set. We then repeated the procedures above on the representative genomes to further subselected 1,478 genomes of <99.3% identities, which were used as the seeds for pan-genome estimation.

We applied PEPPAN [42] to predict a pan-genome of 52,415 genes based on the 1,478 genomes in the seed set. Furthermore, we employed the EToKi MLSTdb module to identify and remove potential paralogs that shared over 80% amino acid similarity. A total of 42,061 pan genes were kept and used to build the whole-genome MLST (wgMLST) scheme for *Klebsiella*. We estimated the presence of genes in the wgMLST scheme in all 33,272 genomes using DTy (https://github.com/ADSGF203com/DTy) and selected a subset of 3,058 core genes that (1) present in ≥95% of genomes, and (2) maintained intact open reading frames in >94% of its alleles using EToKi cgMLST module. The distributed cgMLST scheme was built based on the core genes and made publicly available as part of the KleTy pipeline at https://github.com/zheminzhou/KleTy. DTy designated each allele in the dcgMLST based on the MD5 hash value of its sequence, rather than an arbitrary sequential integer from a central database in the traditional cgMLST scheme. This allowed each genome to be characterized as a collection of up to 3,058 MD5 hash values, which each represented a unique core gene allelic sequence. All the genotyping results were also available in the KleTy repository.

Additionally, we hierarchically grouped the allelic profiles of all genomes into multi-level clusters using pHierCC [29] and statistically evaluated the consistencies and cohesiveness of each cluster using the pHCCeval module, which is also in the pHierCC package.

### Scheme of plasmid clusters (PCs)

The complete sequences of 103,412 plasmids, spanning >2,400 species across the Tree of Life, were downloaded from GenBank (as of March 2023). Sequences that shared high similarity with bacterial chromosomes or viruses (≥95% identities and ≥60% coverages) were identified based on BLAST searches against all complete sequences of bacteria and viruses (https://www.ncbi.nlm.nih.gov/genomes/GenomesGroup.cgi), which were also downloaded from GenBank at the same time. A total of 102,248 high-quality plasmids were retained. We employed BinDash [43] to estimate the pair-wise genetic distances of the plasmids and grouped them into 28,225 single linkage clusters of ≥99% identities. The reference plasmid dataset was built by selecting one sequence of the greatest size for each cluster. Furthermore, we employed FastANI [44] with parameters of “--fragLen 1000 –minFraction 0.5” to calculate average nucleotide identity (ANI) between pairs of plasmids in the reference dataset and used the Leiden algorithm [12] to separate them into 15,797 plasmid clusters (PCs) with ANIs of ≥90% and alignment coverage of ≥50% (Fig. 1a). The similarity network of the plasmids and the resulting PCs were visualized using the Fruchterman Reingold layout algorithm [45] implemented in the Gephi software [46]. The resulting reference sequences, together with the associated host species and PC assignments, were all deposited in the KleTy repository (https://github.com/zheminzhou/KleTy/tree/main/db).

The plasmid prediction module in KleTy employs BLASTn [47] to align each *Klebsiella* genome onto the reference plasmids. To remove potential nonspecific matches, it removes any alignment with <85% identity or <400bp length. Additionally, KleTy evaluates each contig by its alignment coverages to reference plasmids and keeps only alignments that are located in contigs with ≥20% of its sequences similar to the corresponding plasmid. Furthermore, KleTy identifies the PCs iteratively using a greedy algorithm. In each iteration, a PC that has the greatest proportion of its sequences found in the assembly is selected, and all contigs similar to the PC are removed from the following iterations. The program stops when no PCs had >50% of its sequences aligned by contigs, and reports all the identified PCs as well as their associated contigs.

### Plasmid annotation and network construction

Replicon typing and MOB typing of the plasmids were obtained using MOB-typer [10]. Furthermore, the ARGs and VFs were predicted as regions sharing ≥80% identities and ≥60% coverages with reference proteins in the “Bacterial Antimicrobial Resistance Reference Gene Database” (BARRGD) and the hypervirulence genes hosted in Kleborate [4].

We visualized the association between bacterial hosts and the PCs in *Enterobacteriaceae* as a network, in which the nodes represent PCs or bacterial genera. Edges were drawn between a PC and bacteria if the PC was found in the corresponding genus. The resulting network was rendered and visualized using the OpenOrd layout algorithm [48] in Gephi.

### Phylogenetic analysis

The minimum spanning tree of 33,272 *Klebsiella* genomes was constructed using the MSTreeV2 algorithm and visualized in GrapeTree [49]. The supertree of 7,269 representative genomes was estimated using a divide-and-conquer algorithm implemented in the cgMLSA package [11]. The maximum-likelihood trees of HC1360_3, HC1360_8, and the plasmids were all calculated using the EToKi package [38]. Briefly, EToKi employs minimap2 [50] to align all genomes or plasmids onto a reference sequence (Table S9) to obtain a multi-sequence alignment. It also estimates a maximum-likelihood tree based on the alignment using IQ-TREE [51]. We then identified and removed regions in the alignments that were imported by homologous recombination using RecHMM [52] and estimated the tree again based on the remaining non-recombinant regions using IQ-TREE. All resulting trees were visualized online using either GrapeTree or iTOL v6 [53].

### Inferences of population dynamics for HC1360_3

The spatiotemporal dynamics of the HC1360_3 population were estimated by TreeTime [54] based on the maximum-likelihood tree. To calculate the origin dates for the nodes of the tree, we removed all genomes without isolation dates from the tree, and ran TreeTime on the remained tree with the parameters of ‘--keep-polytomies --confidence –covariation --time-marginal always --relax .5 .5 --coalescent skyline --n-skyline 20’. We then run TreeTime again on the dated tree in the ‘mugration’ mode to estimate the ancestral international transmission of the bacteria with the default parameters.

### Statistical analysis

All statistical analyses were performed using R v4.2.2 or Python v3.8. The Cramer’s V statistic was used to test the strength of association between two categorical variables and computed using the rcompanion package in R. The Pearson’s correlations between the carbapenem resistance, geographic distributions, and sizes of HC1360s were performed using the cor.test function in the stats package in R. Linear regression analysis was performed for the carbapenem resistance and geographic distribution of the HC1360s using the ggplot2 package (Method = lm) in R. The similarity of HC1360s and CCs was assessed as the adjusted Rand index (ARI) using scikit-learn in Python. The precision, sensitivity, and F1-score for the results of each plasmid prediction pipeline were also calculated using scikit-learn. A p-value of <0.01 was considered statistically significant in all tests.

## Availability of data and materials

### Data availability

The entire set of 33,272 public *Klebsiella* genomes is available in GenBank, with the accession codes in Table S8. Source data for the figures (including Supplementary Figures) are provided in this paper.

### Code availability

The KleTy command-line toolbox is hosted on GitHub at https://github.com/zheminzhou/KleTy. A detailed README and test dataset are included.

## Acknowledgments

We thank Dr. Karl Drlica for his critical reading and valuable comments.

## Authors’ contributions

Conceptualization: ZZ, YH, and HL; resources: XL, HL, ZZ, and YG; methodology: ZZ, SL, SX, and YR; software: SL, GJ, and XL; writing-original draft preparation: HL and XL; writing-review and editing: HL, XL, JR, LZ, QJ, YH, WS, and ZZ; project administration: ZZ; All authors read and approved the final manuscript.

## Ethics declarations

### Ethics approval and consent to participate

Not applicable.

## Supplementary Figures

**Supplementary Figure 1 |** Proportions of plasmids in each PC with specific Inc types (a) or MOB types (b). Circle size represents the number of plasmids in each PC. The color in the circle indicates the Inc types (a) or MOB types (b) according to the Keys.

**Supplementary Figure 2 |** Proportions of carbapenemase genes carried by each plasmid cluster (PC). Circle size represents the number of plasmids in each PC. The pie chart in the circle indicates the carbapenemase genotypes carried by each plasmid.

**Supplementary Figure 3 |** Dynamic distribution of carbapenemase genotypes in each of five predominant HC1360 populations over the years.

**Supplementary Figure 4 |** The maximum likelihood tree of HC1360_3 (CC147) with branches recalibrated by TreeTime. Dates next to particular branches indicate their estimated dates of origin with the 95% confidence intervals in the brackets. The purple, blue, and green shaded areas indicate clades of ST273, ST392, and ST147, respectively.

## Supplementary Tables

**Supplementary Table 1.** Breakdown of the metadata and ARG genotypes of the HC1360 clusters.

**Supplementary Table 2.** Species and clonal complex of the genomes in each HC1360 cluster.

**Supplementary Table 3.** Metrics to quantify the importance of nodes in the plasmid-exchange network in Figure 2a.

**Supplementary Table 4.** A summary of the performance of plasmid prediction pipelines in the benchmark dataset.

**Supplementary Table 5.** The distribution of genomes, HC1360 clusters, and PCs in *Klebsiella* genus.

**Supplementary Table 6.** Statistics, ARGs, and MOB genotypes of the plasmid clusters (PCs).

**Supplementary Table 7.** Reported bacterial hosts for the newly identified plasmids in *Klebsiella*.

**Supplementary Table 8.** The metadata of 33,272 *Klebsiella* genomes.

**Supplementary Table 9.** Reference genomes used to construct phylogenetic trees.

