## Supplementary Figure 1 for "KleTy: integrated typing scheme for core genome and plasmids reveals repeated emergence of multi-drug resistant epidemic lineages in Klebsiella worldwide"

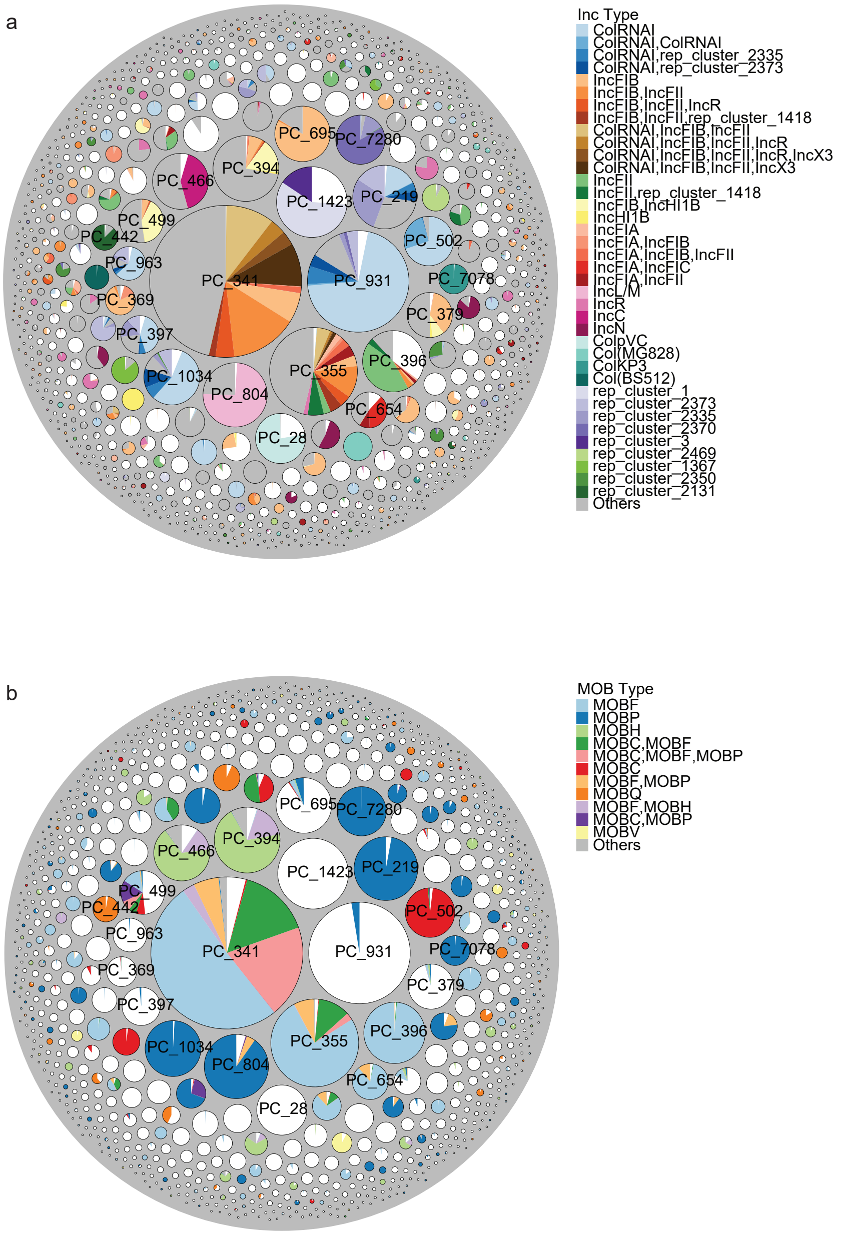


**Supplementary Fig. 1** Proportions of plasmids in each PC with specific Inc types (a) or MOB types (b). Circle size represents the number of plasmids in each PC. The color in the circle indicates the Inc types (a) or MOB types (b) according to the Keys.
