## Supplementary Figure 2 for "KleTy: integrated typing scheme for core genome and plasmids reveals repeated emergence of multi-drug resistant epidemic lineages in Klebsiella worldwide"

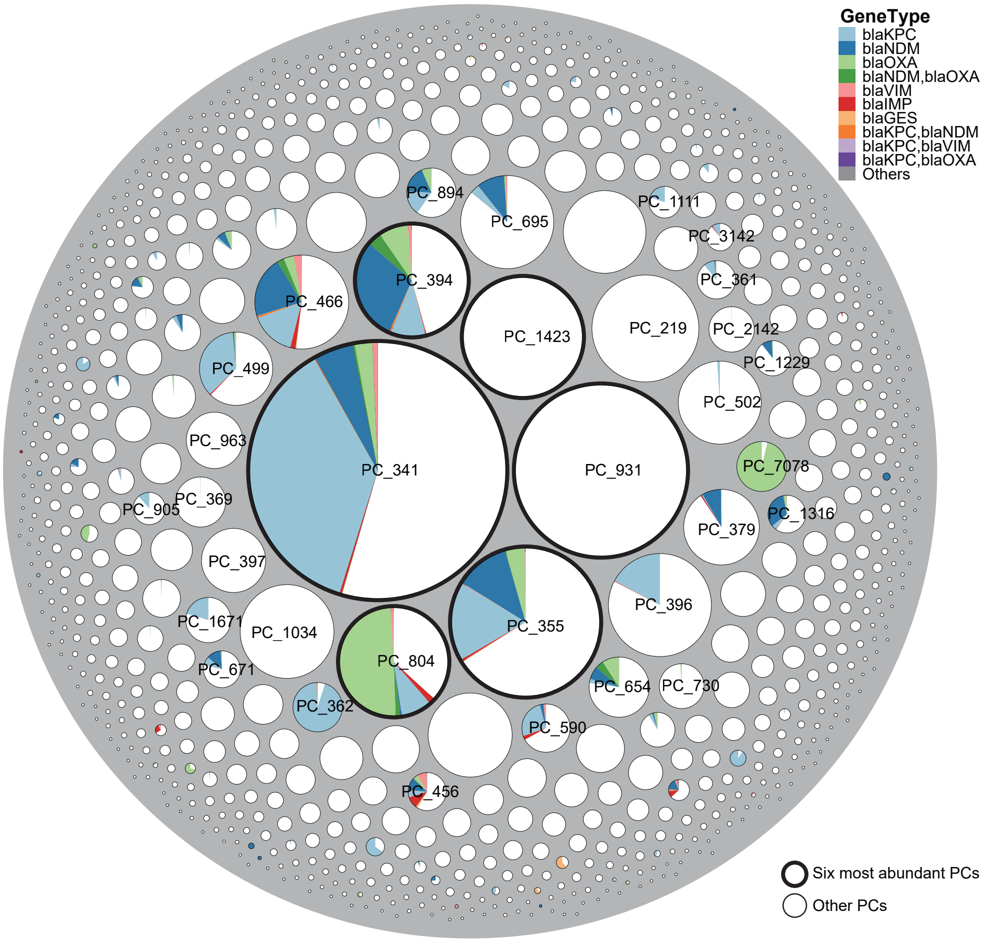


**Supplementary Fig. 2** Proportions of carbapenemase genes carried by each plasmid cluster (PC). Circle size represents the number of plasmids in each PC. The pie chart in the circle indicates the carbapenemase genotypes carried by each plasmid.
