## Supplementary Figure 3 for "KleTy: integrated typing scheme for core genome and plasmids reveals repeated emergence of multi-drug resistant epidemic lineages in Klebsiella worldwide"

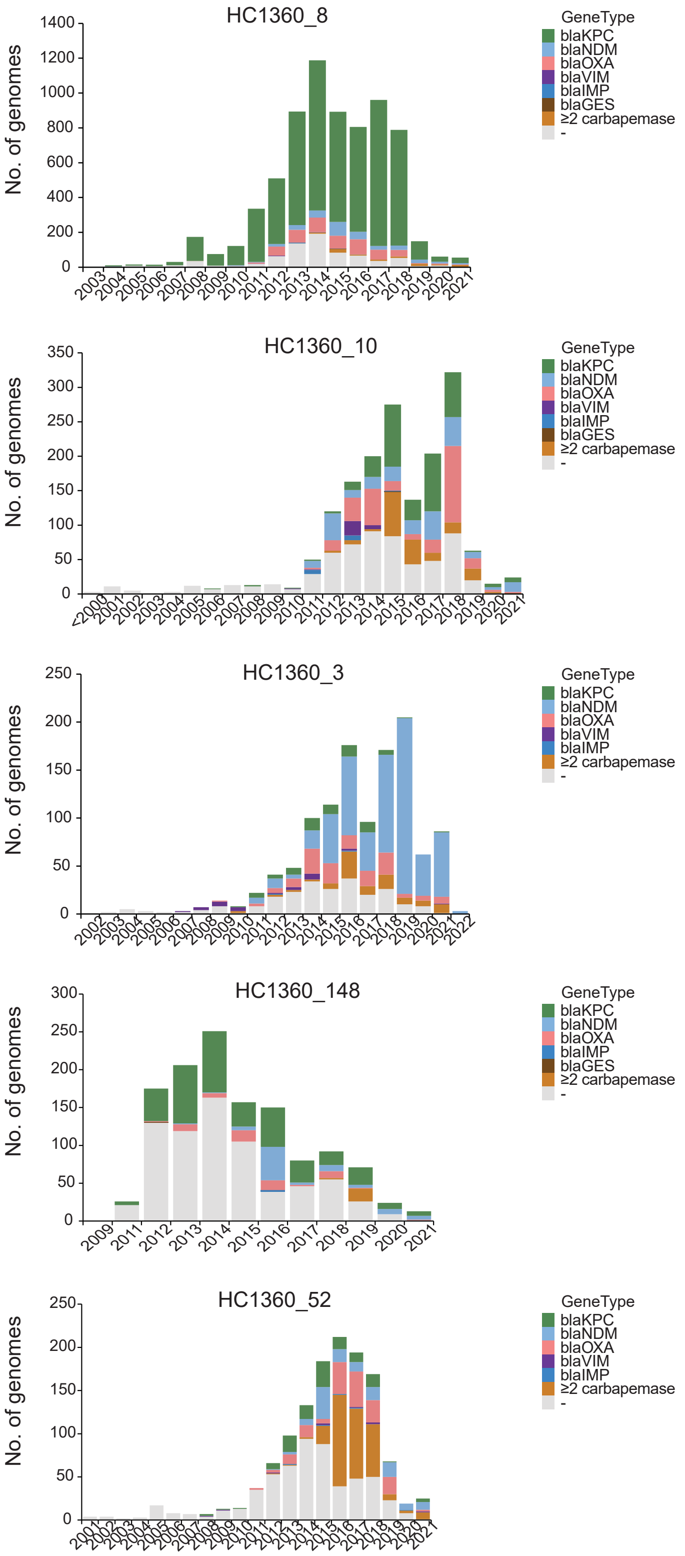


**Supplementary Fig. 3** Dynamic distribution of carbapenemase genotypes in each of five predominant HC1360 populations over the years.
