## Supplementary Figure 4 for "KleTy: integrated typing scheme for core genome and plasmids reveals repeated emergence of multi-drug resistant epidemic lineages in Klebsiella worldwide"

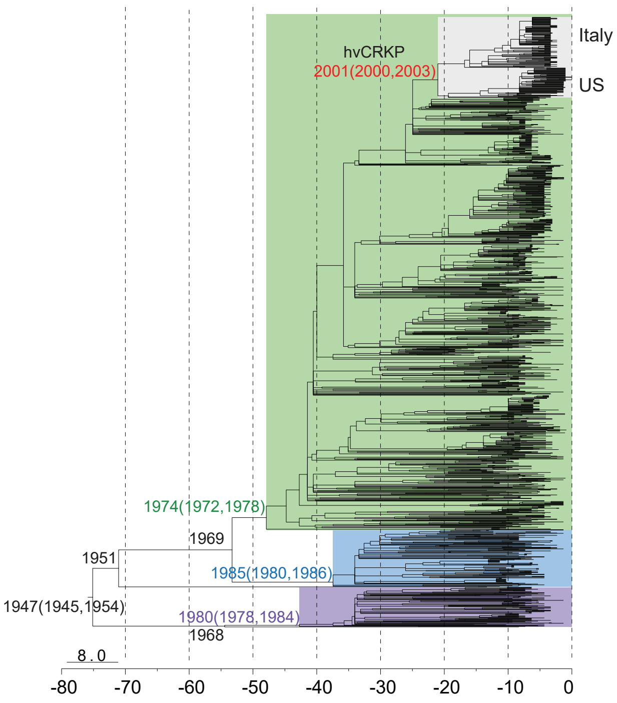


**Supplementary Fig. 4** The maximum likelihood tree of HC1360_3 with branches recalibrated by TreeTime. Dates next to particular branches indicate their estimated dates of origin with the 95% confidence intervals in the brackets. The purple, blue, and green shaded areas indicate clades of ST273, ST392, and ST147, respectively.
